# Characterization of the skin microbiota in bullous pemphigoid patients and controls reveals novel microbial indicators of disease

**DOI:** 10.1101/2021.04.30.21256380

**Authors:** Meriem Belheouane, Britt M. Hermes, Nina Van Beek, Sandrine Benoit, Philippe Bernard, Kossara Drenovska, Sascha Gerdes, Regine Gläser, Matthias Goebeler, Claudia Günther, Anabelle von Georg, Christoph M. Hammers, Maike M. Holtsche, Bernhard Homey, Orsolya N. Horváth, Franziska Hübner, Beke Linnemann, Pascal Joly, Dalma Márton, Aikaterini Patsatsi, Claudia Pföhler, Miklós Sárdy, Laura Huilaja, Snejina Vassileva, Detlef Zillikens, Saleh Ibrahim, Christian D. Sadik, Enno Schmidt, John F. Baines

**Author notes:** **Corresponding author:** John F. Baines, Max Planck Institute for Evolutionary Biology, August-Thienemann-Str. 2, 24306 Plön, Germany. Authors contributed equally to this work (Meriem Belheouane, Britt M. Hermes). Authors contributed equally to this work (Enno Schmidt, John F. Baines).

## Abstract

**Introduction:** Bullous pemphigoid (BP) is the most common autoimmune blistering disease. It predominately afflicts the elderly and is significantly associated with increased mortality. The observation of age-dependent changes in the skin microbiota as well as its involvement in other inflammatory skin disorders suggests that skin microbiota may play a role in the emergence of BP blistering. We hypothesize that changes in microbial diversity associated with BP might occur before the emergence of disease lesions, and thus could represent an early indicator of blistering risk.

**Objectives:** The present study aims to investigate potential relationships between skin microbiota and BP and elaborate on important changes in microbial diversity associated with blistering in BP.

**Methods:** The study consisted of an extensive sampling effort of the skin microbiota in patients with BP and age- and sex-matched controls to analyze whether intra-individual, body site, and/or geographical variation correlate with changes in skin microbial composition in BP and/or blistering status.

**Results:** We find significant differences in the skin microbiota of patients with BP compared to that of controls, and moreover that disease status rather than skin biogeography (body site) governs skin microbiota composition in patients with BP. Our data reveal a discernible transition between normal skin and the skin surrounding BP lesions, which is characterized by a loss of protective microbiota and an increase in sequences matching *Staphylococcus aureus*, a known inflammation-promoting species. Notably, *S. aureus* is ubiquitously associated with BP disease status, regardless of the presence of blisters.

**Conclusion:** The present study suggests *S. aureus* may be a key taxon associated with BP disease status. Importantly, differences in a few key indicator taxa reliably discriminate between patients with BP and matched controls. This may serve as valuable information for assessing blistering risk and treatment outcomes in a clinical setting.

## INTRODUCTION

Bullous pemphigoid (BP) is the most common autoimmune skin blistering disease (AIBD) in Europe, with an annual incidence of about 20 new cases per million in this region [1–3]. It occurs when autoantibodies attack two structural hemidesmosomal proteins of the epidermal basement membrane, i.e. BP180 (type XVII collagen) and BP230, resulting in subepidermal blistering [2,4,5]. The severity of this highly pruritic AIBD considerably affects quality of life and is associated with significantly increased mortality [5]. The incidence of BP is increasing with the aging European population [2,3]. It is thus intriguing that a recent multinational study of 9,000 participants showed that skin microbiota are a predictor of age, more so than oral or gut microbiota [6].

The observation of age-dependent changes in the skin microbiota as well as its involvement in other inflammatory skin disorders suggests that skin microbiota may play a role in the emergence of AIBD. While certain HLA haplotypes are associated with BP, such as HLA-DQA1*05:05 and HLA-DRB1*07:01 in Germans [7], few triggering factors apart from age, medication use, and neuro-psychiatric disease are described [2,8,9]. Previous efforts by Srinivas et al. [10] demonstrated that genotype-dependent microbiota affect disease susceptibility in a mouse model of epidermolysis bullosa acquisita, an AIBD with autoantibodies against type VII collagen. Similarly, Miodovnik et al. [11] presented pilot human data suggesting that skin microbiota contribute to the pathogenesis of BP. However, identification of candidate bacterial taxa or important changes in microbial diversity associated with BP remain uncharacterized.

Here, we conducted a large-scale investigation of patients with BP and age- and sex-matched controls within Europe to clarify relationships between microbiota and BP. By examining skin microbiota surrounding (i) BP lesions, (ii) unaffected skin areas in BP patients, and (iii) controls matched for sex, age, and body site, we reveal clear microbial indicators of both BP disease status and blistering status. The detection of microbial taxa associated with AIBD blistering could enable early intervention and thus, better clinical outcomes.

## MATERIALS AND METHODS

### Study participants

Four-hundred eighteen volunteers were recruited from fourteen study centers across Europe (Germany: Dresden, Düsseldorf, Freiburg, Homburg, Kiel, Lübeck, Munich, Würzburg; France: Reims, Rouen; Sofia, Bulgaria; Thessaloniki, Greece; Oulu, Finland) between October 2015 and September 2019, approved by the University of Lübeck ethics committee (15-051, 18-046), as well as the respective committees of the study centers, following the Declaration of Helsinki. Written, informed consent was obtained from each participant.

Patients with BP were diagnosed according to national and international guidelines and had (i) a compatible clinical picture, (ii) linear deposits of IgG and/or C3 along the dermal-epidermal junction by direct immunofluorescence of a perilesional skin biopsy, and iii) serum IgG reactivity against the epidermal side of human salt-split skin or BP180 NC16A ELISA [12,13]. Patients with BP (n = 228) included 114 males, 113 females, and one sex “unspecified” participant, with an average age of 80 ±8.95 (SD) years (range, 49 to 98 years), and with newly diagnosed or relapsed BP. No newly diagnosed patient had received systemic treatment using dapsone, doxycycline, or immunosuppressants (with the exception of corticosteroids described below) at the time of sampling. All patients had abstained from topical antiseptics two weeks prior to sampling. Systemic and topical corticosteroids had not been administered for BP for longer than 7 days before skin swaps were taken. None of the swabbed individuals received antibiotic therapy, including doxycycline, for at least four weeks.

Age- and sex-matched controls (n = 190) included 104 males and 86 females with non-inflammatory/non-infectious dermatoses with an average age of 80 years ±8.51 (SD) years (range, 47 to 100 years). Controls did not receive systemic antibiotics within four weeks of sampling. Clinical metadata used for the analysis are provided in Supplementary Table S1; summarized demographic and clinical data are provided in Supplementary Table S2.

### Sampling, DNA extraction, and 16S rRNA gene sequencing

Samples were collected using Epicentre Illumina collection swabs (Madison, WI, USA) immersed in 600 uL buffer (50 mM Tris, 1 mM EDTA, 0.5% Tween-20) (Teknova, United States). The swabs were rubbed across the selected body site for 30 seconds and then placed back into the buffer solution. Immediately after swabbing, swabs were stored at -80°C until further processing.

Skin samples (n = 2,956) were obtained from patients with BP representing different cutaneous microenvironments, including “perilesional” skin (defined as being within 2 cm of a primary BP lesion, i.e. a fresh blister or erosion), unaffected skin at the same anatomical location on the contralateral side of the patient (referred to as “contralateral”), and unaffected skin in areas that do not typically manifest disease (we selected the forehead and upper back, as described by Schmidt and Groves [14]), in addition to the antecubital fossa, which was sampled in the human microbiome project [15], collectively referred to as “sites rarely affected by BP”). Two separate perilesional sites from anatomically different BP lesions were sampled from each patient to account for differences in skin biogeography. The locations of lesioned skin, and therefore, perilesional sampling sites, varied from patient to patient. The most common sites included the thigh, arm, foot, knee, lower leg, and hand.

Control participants were swabbed at locations that approximated the sampled body sites in the patients with BP (referred to as “corresponding sites”), in addition to the three sites rarely affected by BP (Figure 1a). Ambient air samples (n = 19), collected by holding a swab in the air for 30 seconds and then placing the collection swab directly into the buffer solution, served as negative sampling controls in addition to negative extraction controls (n = 43). Negative controls were processed alongside samples.

**Figure 1.**
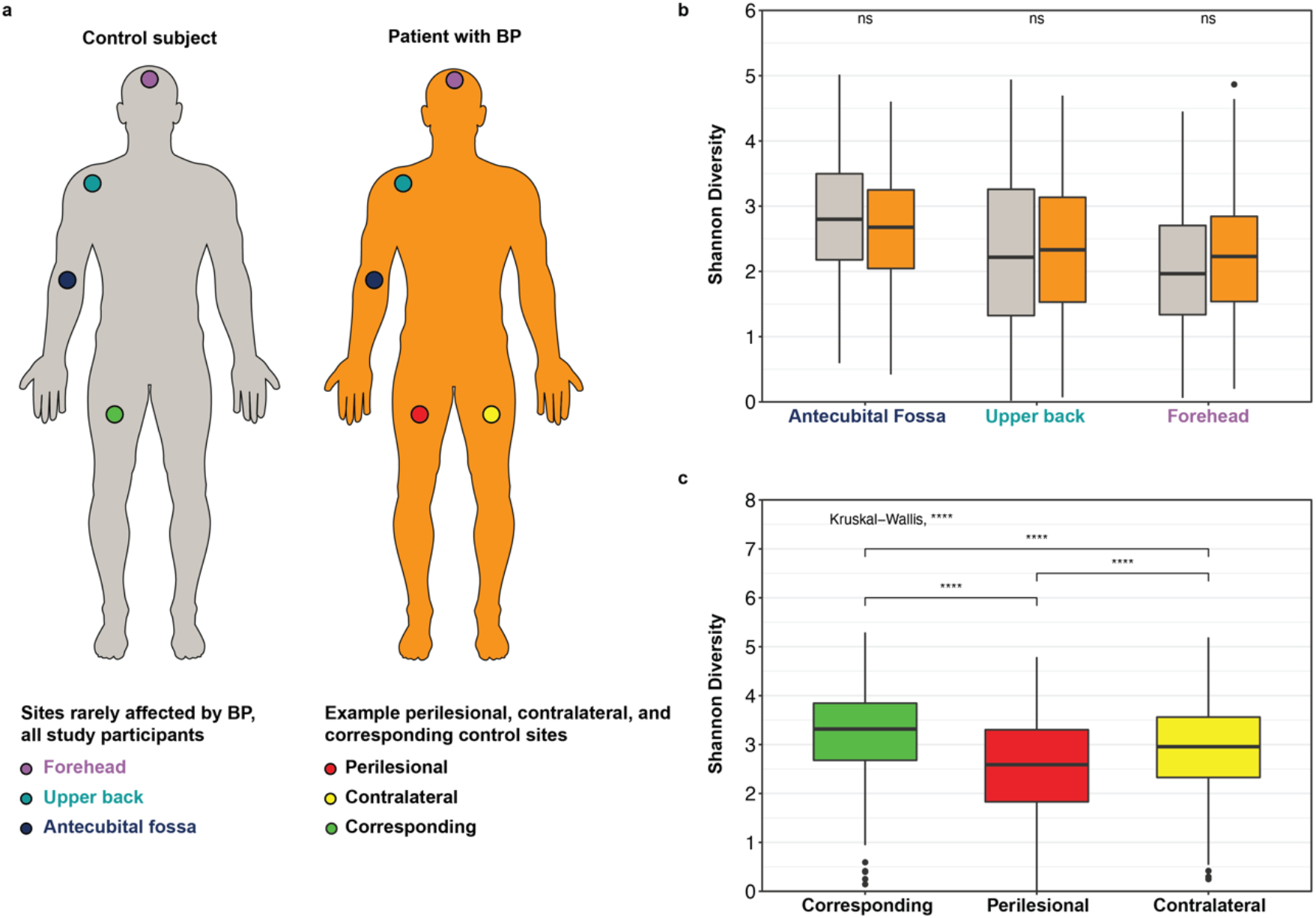
Sampling sites for patients with BP and matched controls; Box plots of Shannon (alpha) diversity. **1a**. Grey figure represents age-and sex-matched control; orange figure represents a patient with BP. Sites rarely affected by BP [see ref. 2] include the forehead (purple), upper back (turquoise), and antecubital fossa (dark blue) are represented on both figures. An example perilesional sampling site (red), unaffected contralateral site (yellow) on the patient, and control-matched corresponding site (green) are shown. **1b**. Shannon diversity at the ASV-level for sites rarely affected by BP for controls and patients. **1c**. Shannon diversity at the ASV-level for patient perilesional, patient contralateral, and control corresponding sites. For box plots: Boxes represent interquartile range between first and third quartiles; horizontal line defines the median. Whiskers represent lowest and highest values. Kruskal-Wallis test applied to analyze site variation. If an overall significant difference was observed, a pairwise Wilcox test was performed; p-values adjusted using the Benjamini-Hochberg method. Significance represented by: * ≤ 0.05; ** ≤ 0.01; *** ≤ 0.001; **** ≤ 0.0001. Supplementary Table S6 reports summary statistics.

ZymoBIOMICS Microbial Community Standard cells (Zymo Research) were used as extraction and sequencing controls to assess contamination in downstream analyses, following the mock community dilution series protocol as described by Karstens et al. [16]. In brief, the strategy is based on the logic that with decreasing “true” microbial biomass (i.e., skin microbes or mock cells), potential signal from background/contamination introduced throughout the procedure will increase. All mock dilutions, as well as the undiluted mock community standards, were treated as samples throughout the extraction, PCR, sequencing, and data processing steps.

Swabs immersed in buffer were thawed overnight at 4°C, then vortexed at high speed for 1 minute. After swab removal, tubes were centrifuged at high speed for 15 min, and the pellets were resuspended in Power Bead solution. DNA was subsequently extracted using the Qiagen DNeasy UltraClean 96 Microbial Kit [96-well plate] (Germantown, MD, USA), according to the manufacturer’s instructions, and eluted in 50uL of the elution buffer. Negative extraction controls were included for each 96-well plate. Samples were stored at -20°C until further processing.

PCR and sequencing were performed by implementing the dual-index sequencing strategy for amplicon sequencing on the MiSeq Illumina sequencing platform, as previously described [17]. Final sample sizes included 2,319 skin swabs comprising 1,451 patient and 868 matched control swabs. A detailed description of sampling methodology and sample processing is provided in Supplementary Methods.

### Data processing and taxonomic classification

The challenges of studying low biomass communities such as skin microbiota are well-documented and include exogenous bacterial DNA contamination from sources such as laboratory reagents, air, and sample collection instruments [16,18–21]. A detailed description of the steps implemented to account for potential contamination is provided in Supplementary Methods. Briefly, data processing and statistical analyses were performed using R (version 4.0.2). Sequences were processed using DADA2 (version 1.16.0), resulting in abundance tables of ASVs [22]. To normalize sequencing coverage, random sub-sampling to 5,000 sequences per sample was performed [23]. Decontam (version 1.8.0; [18] was used within Phyloseq (version 1.32.0; [24] to identify potential contaminant ASVs, according to the prevalence method [16]. ASVs classified to families Halomonadaceae (n = 1,040) and Shewanellaceae (n = 211) were removed, following recommendations of Weyrich et al. [25]. Summary read data are provided in Supplementary Table S3. Taxonomic assignment of ASVs was completed in DADA2 with the Bayesian classifier using the NR Silva database training set, version 138 [26]. Representative 16S rRNA gene sequences were queried via the Ribosomal Database Project (RDP; release 11.6; [27] SeqMatch (version 3; [28]; Supplementary Table S4).

### Ecological and statistical analyses

Analyses included several patient and disease categories. Disease status refers to patients with BP versus matched controls. Blistering status refers to patient perilesional sites versus unaffected, contralateral sites of the same patient. Disease activity was calculated using the Bullous Pemphigoid Disease Area Index (BPDAI). The activity score of both skin and mucosa were combined to account for disease activity, while damage and pruritus points were not considered for calculations. Supplementary Table S5 provides the BPDAI scores for study participants.

Statistical analyses were performed in R (version 4.0.2). Alpha diversity was measured using Shannon and Chao1 indices with vegan (version 2.5-6) on absolute abundance data. Beta diversity was calculated using the Bray-Curtis dissimilarity index. We performed a non-parametric multivariable analysis of variance using distance matrices (PERMANOVA) using the “adonis” function with 1,000 permutations and a partial constrained principal coordinate analysis of beta diversity measures using the “capscale” function in vegan [29]. The significance of models, axes, and terms were assessed using the “anova.cca” function with 1,000 permutations.

Indicator species analysis was applied using indicspecies (version 1.7.9) with the “r.g.” function [30] and 100,000 permutations. Random Forest classification and regression analyses were performed using randomForest (version 4-6-14; [31]. Models were constructed with 100,000 trees, with the “mtry” parameter set for each model and linear models constructed to evaluate potential disease effects. Adjusted R^2^ values reported, beta coefficient values express directionality. Further details are provided in Supplementary Methods.

## RESULTS

### Sampling

Two hundred twenty-eight patients with BP and 190 age- and sex-matched controls from fourteen study sites across Europe were included (see Methods). We performed 16S rRNA gene sequencing on bacterial genomic DNA derived from swabbing four categories of body sites (Figure 1a). These include areas adjacent to a fresh blister or erosion (“perilesional”), non-lesional skin contralateral to the perilesional sample on the same patient (“contralateral”), and the same body site on an age- and sex-matched control (“corresponding”). The locations of the perilesional sites varied from patient to patient. Sites considered to be rarely affected in BP, i.e., forehead and upper back [14], as well as the antecubital fossa, which was sampled in the human microbiome project [15], were sampled in both patients with BP and controls to obtain a more complete picture of the skin microbiota in BP across skin biogeography (Figure 1a).

### Reduced alpha diversity within lesional and BP-susceptible sites

At the ASV-level, we found that the Shannon (Figure 1b) and Chao1 (Supplementary Figure S1a) indices are similar in patients and controls at sites rarely affected by BP. In contrast, control corresponding sites display higher bacterial diversity than patient contralateral sites, which in turn are more diverse than perilesional sites for Shannon (Figure 1c) and Chao1 indices (Supplementary Figure S1b). Supplementary Table S6 provides the summary statistics for group comparisons.

Critically, study center, disease status (i.e., patient with BP versus matched control), and sex significantly correlate with Shannon diversity for patient perilesional and contralateral sites as well as for control corresponding sites (*F* _37,1118_ = 7.24; R^2^_adj_: 0.17; *p < 0*.*001*), with disease status explaining 8.28% of the variance and study center and sex explaining 5.3% and 1.3% of the variance, respectively. Likewise, disease status and study center significantly correlate with Chao1 richness (*F* _37,1118_ = 6.03; R^2^_adj_ = 0.14; *p < 0*.*001*), with study center explaining 7.86% and disease status explaining 1.41% of the Chao1 variance. Disease status associates with a decrease in Shannon diversity in patient perilesional and contralateral sites (β = -0.72, -0.38, respectively), and a decrease in Chao1 richness (β = -39.38, -30.90, respectively). Thus, disease status associates with a substantial decrease in alpha diversity, which is still present after accounting for potential confounding variables. To determine whether these findings were affected by spatial correlation across body sites, we calculated a linear mixed model using “individual” as a random term to estimate variability in alpha diversity measures and to control for non-disease variables, including sex and study center. The model reveals statistically significant variance similar to that estimated by the above linear models, suggesting that the results reported here are unlikely to be conflated by cases of multiple measures of diversity (see Supplementary Results).

Analysis of sum of squares shows that the effect of disease status on alpha diversity does not extend to sites rarely affected by BP. Rather, skin biogeography likely characterizes microbial diversity at these sites (see Supplementary Results). Of note, the severity of disease as determined by the Bullous Pemphigoid Disease Area Index (BPDAI) [32], did not significantly associate with mean alpha diversity measures at perilesional and contralateral skin sites.

### Beta diversity in relation to disease, individual, and sampling features

We first analyzed beta (between-sample) diversity at sites rarely affected by BP to evaluate the effects of potential confounding variables (see Supplementary Methods). Analysis of disease status per body site reveals a significant association with disease status (adonis: Bray-Curtis∼disease status: body site; R^2^=0.003; *p* < 0.001). Partial constrained principal coordinate analysis reveals that patients and controls cluster according to body site along the first and second axes and that forehead and upper back (typical sebaceous zones) are more similar to each other compared to the antecubital fossa (Figure 2b). These findings suggest that the microbial variation among sites rarely affected by BP is likely linked to skin biogeography rather than disease or study center (Figure 2a, 2c). Additionally, partial constrained principal coordinate analysis reveals that on the first and second axes, control corresponding sites are distinguishable from patient contralateral and perilesional sites, which largely cluster together (Figure 2d, e). Figure 2f, on the other hand, shows comparatively little clustering according to study center.

**Figure 2.**
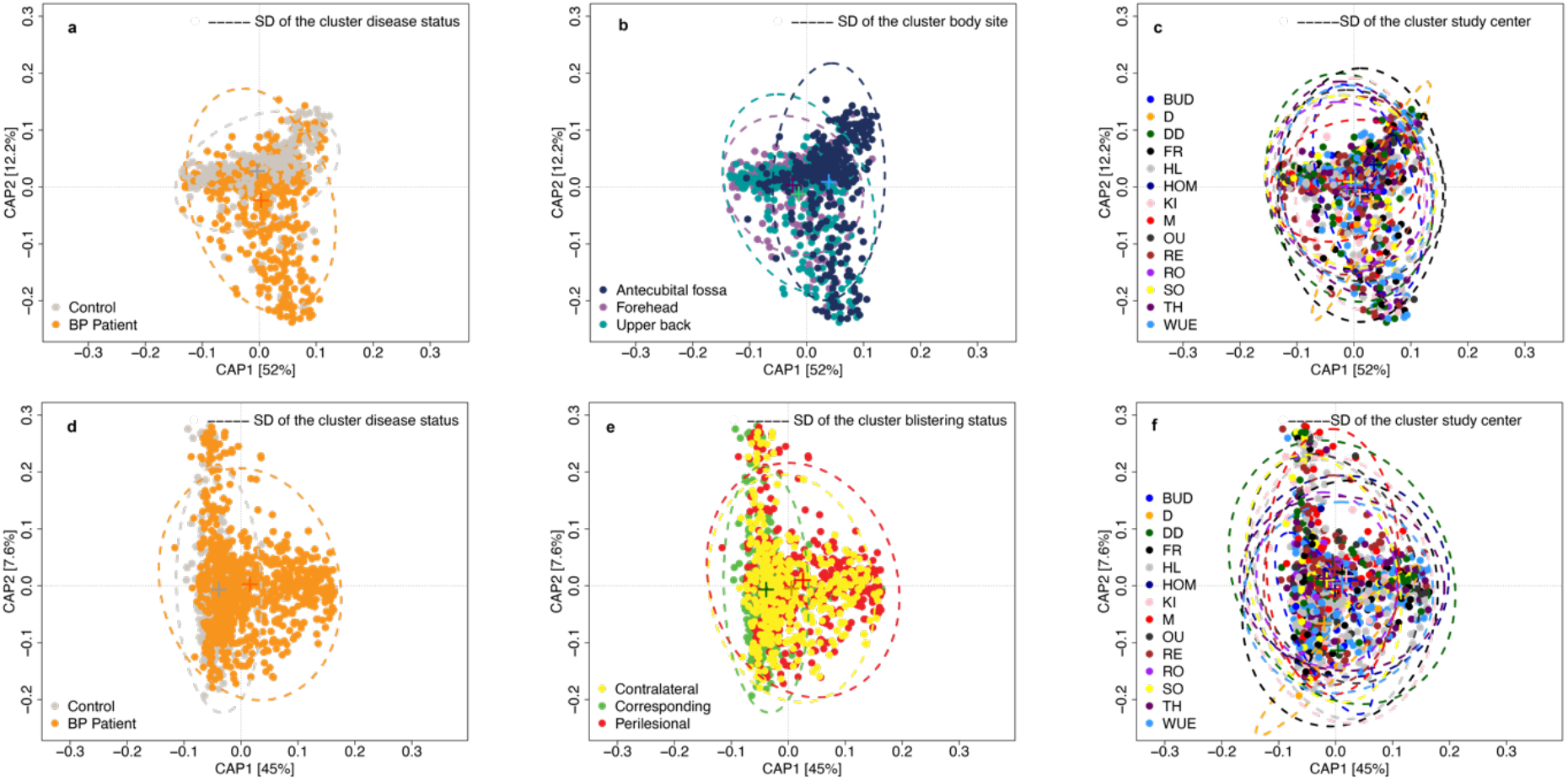
Partial constrained principal coordinate analyses of Bray-Curtis. **2a to 2c**. Body sites rarely affected by BP. (anova.cca, Full model: *p* = 0.0009; terms: disease status, body site (constrained inertia=5.04%, conditioned inertia=4.5%), study center: *p* < 0.001; axes: CAP1, CAP2: *p* = 0.09; 1,000 permutations). **2d to 2f**. Patient perilesional, patient contralateral sites, and control corresponding sites. (anova.cca, Full model: *p* < 0.001; terms: disease status, blistering status, study center: *p < 0*.*001*; axes: CAP1, CAP2: *p* = 0.009; 1,000 permutations). “+” represents centroid. SD: standard deviation. Site abbreviations: Budapest, Hungary (BUD); Düsseldorf, (D), Dresden, (DD), Freiburg, (FR), Lübeck, (HL), Homburg, (HOM), Kiel, (KI), München, (M), Würzburg (WUE), all Germany; Oulu, Finland (OU); Reims, (RE), Rouen, (RO), both France, Sofia, Bulgaria (SO), Thessaloniki, Greece, (TH).

We additionally analyzed beta diversity between patient perilesional, patient contralateral, and control corresponding sites as described above. We find that disease, blistering status, and study center all explain a portion of the variance in beta diversity (see Supplementary Results). However, an analysis of interaction between variables reveals that disease status accounts for significant differences between study centers (adonis: disease status: study center, R^2^ = 0.03; *p*

< 0.001). Furthermore, linear modeling shows that BPDAI significantly correlates with study center (F_13,196_=3.31, R^2^_adj_=0.117, *p* < 0.001). These results suggest that variation between study centers could be explained by differences in patient populations between study centers, e.g., perhaps only the most severely affected patients are seen at university study centers in some regions.

### Indicator species of BP patients and controls

We conducted four indicator species analyses at the ASV-level. To refine the taxonomic classification of indicator ASVs, we queried representative sequences using RDP SeqMatch (see Supplementary Table S4). ASVs strongly associated with BP patients or controls are shown in Figure 3 and Supplementary Table S7.

**Figure 3.**
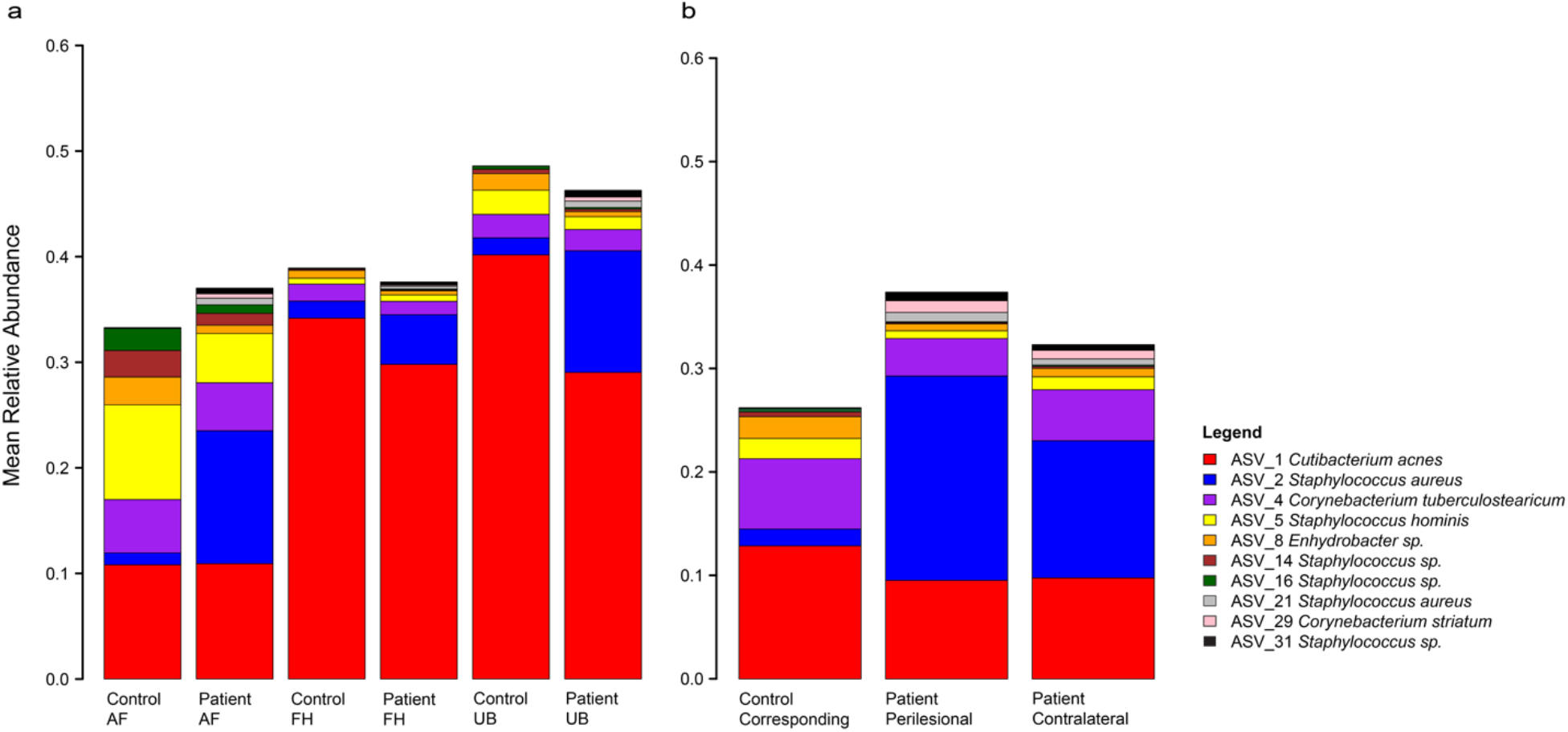
Bar plots of mean relative abundance for the ten most important indicator species. **3a**. Bar plot showing relative abundance of important indicator species, at the ASV-level, for controls and patients with BP at sites rarely affected by BP [antecubital fossa (AF), forehead (FH), and upper back (UB)]. **3b**. Bar plot showing the relative abundances of important indicator species at the ASV-level for patient perilesional, patient contralateral sites, and control-matched corresponding sites. RDP SeqMatch results for the representative ASV sequences are shown in the legend and provided in full in Supplementary Table S4. Supplementary Tables S7, S8 provide statistical parameters for indicator species analyses and summary statistics of all indicator ASVs, respectively.

Several indicator ASVs known to be human commensals associate with sites rarely affected by BP (i.e., forehead, upper back, and antecubital fossa in our study). Importantly, we identify a greater number of ASVs associating with these standardized control sites, which is coherent with the observed loss of diversity in patients with BP. ASV_1, which closely matches *Cutibacterium acnes* (*C. acnes*) [previously known as *Propionibacterium acnes;* [33], is an indicator at the forehead, upper back, and antecubital fossa. Specifically, among these sites rarely affected by BP, body site is associated with 13% of the variance in *C. acnes* abundance (abundance is accordingly higher at the sebaceous forehead and upper back sites), whereas disease status accounts for 0.79% variance of *C. acnes* abundance (*F*_39,1123_ = 11.58; R^2^_adj_ = 0.26; *p < 0*.*001*). Interestingly, disease status is associated with a decrease in *C. acnes* abundance at the upper back and forehead (β = –0.11 and β = –0.045, respectively).

Important patterns are also apparent among control corresponding, patient perilesional, and patient contralateral sites. Within these sites, *C. acnes* abundance associates with study center, blistering status, and sex (*F*_37,1118_ = 5.40; R^2^_adj_ = 0.12, *p < 0*.*001*), with sex explaining 7.17%, study center explaining 4.66%, and blistering status (i.e., patient perilesional versus patient contralateral) explaining 0.63% of the variance. Furthermore, *C. acnes* relative abundance is greater at control corresponding sites and relatively lower at patient perilesional sites (β = +0.03, –0.003, respectively). The higher relative abundance of *C. acnes* at these control corresponding sites is consistent with the increased abundance of *C. acnes* observed in rarely affected sites. Additionally, ASV_4 [which closely matches *Corynebacterium tuberculostearicum* (*C. tuberculostearicum*)] is an indicator for control corresponding sites and patient contralateral sites. Study center, blistering status, and sex are significantly associated with *C. tuberculostearicum* abundance (*F*_37,1188_ = 3.92; R^2^_adj_ = 0.09; *p < 0*.*001*), with blistering status explaining 4.06% of the variance and correlating with an increase in abundance in control corresponding sites, but a decrease in patient perilesional sites (β = +0.05, –0.04, respectively). Summary statistics for indicator ASVs are provided in Supplementary Table S8.

### Contrasting patterns of *Staphylococcus* ASVs in patients with BP and controls

Six indicator ASVs belong to *Staphylococcus* and display contrasting patterns associated with disease status. *Staphylococcus* ASV_5 (which closely matches *S. hominis*) abundance significantly correlates with both disease status and body site (*F*_39,1123_ = 6.45; R^2^_adj_ = 0.16; *p < 0*.*001*). However, as with other indicator ASVs known to be human commensals, body site explains a greater proportion of variance (11.17%) compared to disease status (1.24%), whereby the latter is associated with a decrease in abundance (β = –0.04). Accordingly, *S. hominis* is significantly negatively correlated with BPDAI at patient contralateral sites (Spearman’s rho = -0.17; *p* < 0.05), but there is no relationship between BPDAI and *S. hominis* at patient perilesional sites. In contrast, *Staphylococcus* ASV_2 (which closely matches *S. aureus*) is a strong indicator for BP, including at patient body sites rarely affected by BP. Here, disease status explains 7.35% of the variance in *S. aureus* abundance, whereas body site explains 1.52% (*F*_39,1123_ = 5.12; R^2^_adj_ = 0.12; *p < 0*.*001*). Notably, disease status associates with an increase in *S. aureus* abundance at these rarely affected sites (β = 0.08). However, among perilesional, contralateral, and corresponding sites, blistering status accounts for the greatest amount of variance for *S. aureus* abundance (11.67%), followed by study center (6.45%), and sex (0.77%; *F*_37,1118_ = 9.09; R^2^_adj_ = 0.21, *p < 0*.*001*). This is characterized by a decrease of *S. aureus* abundance in control corresponding sites (β = –0.12) compared to an increase at patient perilesional sites (β = +0.07). Additionally, *S. aureus* positively correlates with BPDAI at perilesional and contralateral sites (Spearman’s rho = 0.2; *p* < 0.01; Spearman’s rho = 0.28; *p* <0.001, respectively; Supplementary Figure S2). To address concerns of spatial correlation across body sites as a potential confounding factor, we constructed a linear mixed model using “individual” as a random term to control for non-disease variables and to estimate variability in mean ASV indicator abundances explained by BPDAI for patients with BP. Estimates reveal similar findings in terms of significance and proportions of variance explained by disease status, except for ASV_1, *C. acnes*, which is not affected by blistering status using this model (Supplementary Results).

Because individual members of *Staphylococcus* can display antagonistic interactions in the context of inflammatory skin disorders [34], we examined pairwise correlations among the top ten indicator ASVs (Supplementary Figures S3, S4; Supplementary Table S9). Importantly, *Staphylococcus* ASV_2 (*S. aureus*) and ASV_5 (*S. hominis*) display significant negative correlations within patient perilesional sites, patient contralateral sites, and at the antecubital fossa site in patients with BP. However, there is no significant correlation between these two *Staphylococcus* indicators at any matched control sites. Furthermore, *Staphylococcus* ASV_2 is significantly negatively correlated with sequences matching *C. acnes* (ASV_1) at all sampling category sites in patients with BP. This association is absent at all sampling category sites in matched controls. This finding suggests a fundamental alteration in community interactions among members of *Staphylococcus* in the context of BP.

### *Staphylococcus* ASVs predict disease status in random forest classification

Random forest classification analyses reveal indicator ASVs to accurately classify samples when applied to all sampling sites (mtry = 15; 849/868 controls and 1,443/1,451 BP patients; mean classification accuracy 99.00%). Prediction accuracy approaches 100% when applied to only control corresponding, patient contralateral, and patient perilesional sites (mtry = 18; 324/334 controls and 822/822 BP patients; mean classification accuracy 99.15%; Supplementary Figures S5a, S5b). By inspecting the mean decrease accuracy components for ASVs, those belonging to the *Staphylococcus* genus are identified as being most important to both models (see Supplementary Table S10).

To estimate the discriminatory power of *Staphylococcus* ASVs alone, we limited the random forest classification analyses to *Staphylococcus* ASVs with an abundance greater than 2% within each sample. We found that *Staphylococcus* ASVs accurately distinguish between controls and patients with BP (mtry = 52; 790/868 controls and 1,446/1,451 patients; mean classification accuracy 96.40%) when applied to all sampling sites and are similarly accurate when applied using only control corresponding, patient perilesional, and patient contralateral sites (mtry = 62; controls 294/334 and 819/822 patients; 96.20%; Supplementary Figures S5c, S5d). Notably, inspection of mean decrease accuracy components indicates that *S. aureus* ASV_2 is the most important ASV for model accuracy.

## DISCUSSION

This study reveals marked differences in the skin microbiota of patients with BP compared to that of sex- and age-matched controls with non-inflammatory/ non-infectious dermatoses. This was accomplished by conducting large-scale sampling and bacterial 16S rRNA gene analysis, utilizing a sampling scheme that accounts for both skin biogeography and disease status (Figure 1a). This study represents the most substantial sampling effort of skin microbiota in BP to date.

We observe a significant reduction in alpha diversity at both perilesional sites and contralateral sites in BP patients compared to site-matched areas from controls. Furthermore, blistering status (i.e., patient perilesional versus unaffected, contralateral sites within patients), as well as disease status, are associated with a fewer number of indicator ASVs when compared to matched corresponding sites from control subjects. This reduction in alpha diversity in patients with BP is consistent with findings from other studies of inflammatory skin diseases, including psoriasis [35,36], atopic dermatitis [37], as well as a mouse model of the BP-like variant epidermolysis bullosa acquisita [10].

The clear biogeography of human skin microbiota, whereby distinct assemblages colonize different body sites depending upon numerous factors, suggests that conditions like BP that affect the skin micro-environment, and thereby skin microbiota, may influence susceptibility to blistering [19,38]. Our data reveal that BP might contribute to a loss of protective microbiota in sites rarely affected by BP. At the upper back, an interaction model revealed that disease status significantly associates with a decrease in *C. acnes* relative abundance in patients with BP. This is notable given that the upper back represents a sebaceous skin zone where we would expect relatively high amounts of *C. acnes* [39,40]. Although *C. acnes* is commonly thought of as a potential pathogenic species responsible for acne, it also acts as an important commensal that aids in preventing the colonization and invasion of pathogens via the production of antimicrobials and hydrolysis of triglycerides [39,41], as well as the production of short-chain fatty acids [42,43]. For *S. hominis*, another human commensal, we also found that disease status associates with a decrease in abundance at rarely affected sites. Furthermore, we found a negative association between disease activity (measured by the validated disease score BPDAI [32]) and *S. hominis* in contralateral sites of patients with BP. In contrast, *S. aureus* relative abundance positively associates with BPDAI scores at perilesional and contralateral sites. Additionally, our data show that in the skin sites of matched controls that correspond to the perilesional sites in patients with BP, there is a relative increase in abundance of the commensals *C. acnes* and *C. tuberculostearicum*. Furthermore, *S. hominis* is relatively decreased in patient contralateral sites, suggesting that the effect of disease extends beyond perilesional sites in patients with BP. It is thus possible that protective effects provided by different commensal bacteria may be fundamentally altered in patients with BP, e.g. the production of antimicrobials by *Staphylococcus* strains, as in atopic dermatitis [34,44]. Decreased commensal microbiota perhaps translate to fewer protective immune functions in the skin, which in turn could allow for increased colonization of inflammation-promoting species like *S. aureus* [34]. Thus, our observations might be capturing a baseline state of disease at sites without blisters, whereby beneficial taxa such as *S. hominis* and *C. acnes* are lost throughout the pathogenesis of the disease.

Accordingly, *S. aureus* is known to dominate the skin microbiota of patients with atopic dermatitis and exacerbates the disease through inflammation [34,45]. Notably, we found that *S. aureus* is associated with disease status regardless of sampling site, suggesting that this taxon is an important indicator of BP. The mean relative abundance of *S. aureus* is increased in sites rarely affected by BP as well as in the perilesional and contralateral sites. These results are in line with a recent study showing BP lesions are frequently colonized by TSST-1^+^ *S. aureus* in patients with new onset disease compared to age- and sex-matched controls [46]. The authors further observed that antibiotic therapy eliminated *S. aureus* and improved clinical outcomes. The specific role of this microbe and its functional components, e.g., how it might drive blister formation, will require further exploration.

In addition to cutaneous micro-environmental differences, geographic locations of patients with BP should be considered, as there is significant global variation in microbial colonization, especially as it relates to disease susceptibility [47–50]. Population differences observed in the gut microbiota in patients with inflammatory bowel disease, for example, suggest a complex interplay between geography and gut diseases that are in part driven by microbial factors [50]. Therefore, a broad-scale sampling of patients with BP across regions with variable incidences could reveal population-specific characteristics that might affect disease predisposition. We found that BPDAI scores explained a portion of microbial taxon variation between study centers. We recognize that geography represents an assemblage of factors including diet, culture, ancestry, and environmental features. Our results suggest the need for a large, global study to disentangle the relative importance of these features on the assembly of the skin microbiota, especially as it pertains to disease onset in AIBD.

## CONCLUSIONS

In summary, our study suggests that skin microbiota may play an important role in the emergence of BP skin lesions, perhaps via the loss of beneficial taxa such as *S. hominis* and/or via the colonization of inflammation producing taxa such as *S. aureus*. Given the clear discriminatory power provided by differences in a few key indicator taxa, their relative proportions have the potential to provide critical information for assessing blistering risk as well as treatment outcomes. Future research may focus on functional analysis of host-microbe and microbe-microbe interactions as a means to identify novel treatment approaches for BP.

## Data Availability

Datasets related to this article can be found under BioProject accession number PRJNA715468 at https://www.ncbi.nlm.nih.gov/bioproject/, an open-source online data repository hosted at the NCBI SRA BioProject database.

https://dataview.ncbi.nlm.nih.gov/object/PRJNA715468?reviewer=tpob0l00b03qg88ju4lqnp7dou

## Abbreviations

AI: autoimmune
BP: bullous pemphigoid
AIBD: Autoimmune blistering disease
(BPDAI): Bullous Pemphigoid Disease Area Index
DF: degrees of freedom
IS: indicator species
ASV: amplicon sequence variant

## Funding sources

This work was supported by the German Research Foundation (DFG), through Clinical Research Unit 303, project number 269234613, subproject P2, jointly awarded to Prof. Dr. John F. Baines and Prof. Dr. Dr. Enno Schmidt.

## DATA AVAILABILITY

Reviewer link: https://dataview.ncbi.nlm.nih.gov/object/PRJNA715468?reviewer=tpob0l00b03qg88ju4lqnp7dou

## ACKNOWLEDGEMENTS

We would like to thank Jan Schubert, Katja Cloppenborg-Schmidt, and Olga Eitel for their excellent technical assistance. We are grateful to Sarah Gaugel and Stephanie Freyher, Lübeck, for technical assistance with sample storage. We are indebted to Ana Luiza Lima, Kaan Yilmaz, and Onur Dikmen, Lübeck, for assistance with sample storage and communication with study centers during various phases of the study.

## CONFLICTS OF INTEREST

The authors state no conflict of interest.

## AUTHOR CONTRIBUTIONS

**Conceptualization**: JFB, ES, SI, CDS; **Data curation:** MB, BMH; **Formal analysis:** MB, BMH; **Funding acquisition:** JFB, ES, CDS; **Investigation:** MB, BMH; **Methodology:** JFB, ES, CDS; **Project administration:** JFB, ES, NVB, MB, CMH, CDS, MMH; **Resources:** MB, NVB, SB, PB, KD, SG, RG, MG, CG, AVG, CMH, MMH, BH, FH, BL, PJ, AP, CP, MS, LH, SV, CDS, ES; **Software:** MB, BMH; **Supervision:** JFB, ES, MB; **Validation:** JFB, MB; **Visualization:** JFB, MB, BMH; **Writing–Original draft preparation:** JFB, BMH, MB; **Writing–Review and editing:** JFB, BMH, MB, NVB, SB, PB, KD, SG, RG, MG, CG, AVG, CMH, MMH, BH, FH, BL, PJ, AP, CP, MS, LH, SV, CDS, ES

## Notes

### Competing Interest Statement

The authors have declared no competing interest.

### Author Declarations

This study was approved by the University of Luebeck ethics committee (15-051, 18-046), as well as the respective committees of the study centers, following the Declaration of Helsinki.

### Summary of Updates

Manuscript updated; Supplemental files updated

