## Supplementary information for "Characterization of the skin microbiota in bullous pemphigoid patients and controls reveals novel microbial indicators of disease"

**SUPPLEMENTARY MATERIALS**

- Supplementary Material Methods
- Supplementary Results: Alpha diversity results not reported in main text
- Supplementary Results: Beta diversity results not reported in main text
- Supplementary Results: Linear mixed model results
- Supplementary Figure S1. Chao1 alpha diversity plots
- Supplementary Figure S2. *S. aureus* correlation with BPDAI
- Supplementary Figure S3. Correlation matrices for patient perilesional, patient contralateral, and control corresponding sites
- Supplementary Figure S4. Correlation matrices for sites rarely involved in BP from patient and control cohorts
- Supplementary Figure S5. Multidimensional plots of the proximity matrix by randomForest

**SUPPLEMENTARY TABLES, SEPARATE EXCEL FILE**

- Supplementary Table S1. Study metadata
- Supplementary Table S2. Summarized demographic and clinical metadata
- Supplementary Table S3. Summarized read counts
- Supplementary Table S4. RDP SeqMatch scores (S_ab) for 370 representative ASV sequences
- Supplementary Table S5. BPDAI disease scores
- Supplementary Table S6. Alpha diversity summary statistics
- Supplementary Table S7. Indicator species for sampling sites (order based on stat parameter)
- Supplementary Table S8. Summary statistics of all significant indicator ASVs (n = 370)
- Supplementary Table S9. Important components of random forest models
- Supplementary Table S10. Correlation coefficients for Spearman's correlation matrices

### SUPPLEMENTARY MATERIAL METHODS

#### 16S rRNA sequencing

Hypervariable regions V1-V2 of the bacterial 16S rRNA gene were amplified. The primer pair 5′-AATGATACGGCGACCACCGAGATCTACACXXXXXXXXTATGGTAATTGT AGAGTTTGATCCTGGCTCAG -3′ and 5′-CAAGCAGAAGACGGCATACGAGATXXXXXXXXAGTCAGTCAGCC TGCTGCCTCCCGTAGGAGT-3′ contained the Illumina P5 (forward) and P7 (reverse) sequences (denoted by italics) and the universal primer 27F and 338R sequences (denoted by underlined italics), which represent broadly conserved regions of the bacterial 16S gene. Per Illumina's recommendations, a twelve-base linker sequence (underlined only) was added to increase the annealing temperature of the sequencing primer. Both primers contained a unique eight base multiplex identifier (designated by XXXXXXXX) to tag each PCR product. PCR was conducted in a 25-uL volume containing 2uL DNA using Phusion Hot Start II DNA High-Fidelity DNA Polymerase (Finnzymes, Espoo, Finland). The cycling conditions were as follows: initial denaturation for 30 seconds at 98°C; 30 cycles of 9 seconds at 98°C, 60 seconds at 50°C and 90 seconds at 72°C; final extension for 10 minutes at 72°C. PCR products were loaded on a 1.5% agarose gel to confirm and quantify the 16S rRNA gene bands. PCR product concentrations were first quantified on an agarose gel using image analysis software (Bio-Rad). After quantification, products were combined accordingly to make equimolar subpools. The subpools were then extracted from an agarose gel using the Qiagen MinElute Gel Extraction Kit and quantified with the Quant-iTTM dsDNA BR Assay Kit on a Qubit fluorometer (Invitrogen). Finally, subpools were combined into one equimolar pool for each library. Pools were further purified using AMPure® Beads (Agencourt), and libraries were run on an Agilent Bioanalyzer prior to sequencing, as recommended by Illumina. The libraries were sequenced on a MiSeq using the MiSeq Reagent Kit v3 600 cycle sequencing chemistry (Illumina, CA, USA).

**Data processing and taxonomic classification**

Raw fastq sequencing reads were generated and demultiplexed to allow for no mismatches in the index sequences (Bcl2fastq, Illumina). Data processing was performed in the R software environment (version 4.0.2) using the DADA2 (version 1.16.0; [1] workflow resulting in abundance tables of amplicon sequence variants (ASVs). All sequencing runs underwent quality control and error profiling separately. Forward and reverse reads were trimmed to a length of 250 and 200 bp, respectively, using standard filtering parameters. Low quality read-pairs were discarded when the estimated error in one of the reads exceeded 2 or if ambiguous bases ("N"s) were present in the base sequence. Read pairs that could not be merged due to insufficient overlap or mismatches in their nucleotide sequences were discarded. All data from the independent sequencing runs were finally combined into a single abundance table, and then underwent chimera removal in DADA2. Taxonomic assignment of ASVs was completed in DADA2 with the Bayesian classifier using NR Silva database training set version 138 [2]. Merged reads that were less than 270 bp or more than 330 bp were excluded. ASVs classified as eukaryotic were excluded.

The Decontam package (version 1.8.0; [3]) was used within Phyloseq (version 1.32.0; ref) to identify potential contaminant ASVs. The prevalence method, which uses presence/ absence information of sequence features for all samples to compare the prevalence of ASVs in negative controls compared to true samples, was used with the strict probability threshold of 0.5 [4]. ASVs tagged as probable contaminants were removed from the abundance table. Furthermore, and as suggested by Karstens et al., an additional filtering step was performed as follows: ASVs present/detected in negative controls (sampling ambient air and extraction controls) for a respective sample that harbored an absolute abundance of less than or equal to 2 percent were excluded from the skin sample, allowing for potential cross-contamination between samples to be accounted for (i.e. true ASVs are less likely to be *falsely* labeled as a contaminant if their abundance is greater than 2 percent). All ASVs classified as belonging to the families Halomonadaceae (n = 1,040) and Shewanellaceae (n = 211) were removed, as these bacteria represent common contaminants in low biomass samples [5].

Following these steps, we verified our methods by performing qPCR on a subset of randomly selected skin samples as well as mock community samples that were serially diluted as described by Karstens et al. [4] to assess the increase of contamination that tends to occur proportionally to decreasing DNA concentration. To define the total bacterial load, we performed qPCR with 3 technical replicates on 49 skin swabs, 43 mock dilutions, 6 negative extraction controls, and 1 air sampling control using Eub338F (ACTCCTACGGGAGGCAGCAG), and Eub518R (ATTACCGCGGCTGCTGG). PCR protocol was as follows: Primer mix 10 uM (0.5 ul), SYBR™ Green PCR Master Mix (5 ul) (Applied Biosystems), Nucleic acid-free water (2.5 ul), and 2ul of cDNA. PCR program: Initial 95°C for 10 min, (denature at 95°C for 15 seconds, anneal and extend at 60°C for 60 sec) for 40 cycles; extend at 60°C for 30 sec, melt ramp at 60-95°C and finish at 4°C for 10 sec.

First, we found that the bacterial DNA concentration, as expressed by Cq values, of our skin samples were similar to the mock community samples with bacterial DNA concentrations serially diluted three to four times (named D3 and D4, respectively). Furthermore, these D3 and D4 samples contained only a small percentage of contamination both before and after implementing the above-described contamination steps, suggesting that samples with similar Cq values likely also contained only a small percentage of contamination, if any. Finally, we demonstrated a strong positive correlation between ASVs classified as contaminates by the Decontam package and Cq values (Spearman rho = 0.80; $p < 0.00001$). None of the best indicator ASVs reported in Figure 3 of the main text correlated with Cq values.

**Ecological and statistical analyses**

Alpha diversity (within sample diversity) was measured using Shannon and Chao1 indices, which account for species richness and evenness, respectively, in vegan (version 2.5-6) on absolute abundances data. Groups were compared using Kruskal-Wallis and pairwise Wilcoxon rank sum tests. Correction for multiple testing was performed according to Benjamini and Hochberg method [6]. To determine whether these findings were affected by spatial correlation across body sites, we fitted a linear mixed effects where blistering status (perilesional, contralateral, or control corresponding) is defined as the fixed effect and individual ID is defined as a random term. The model was fitted by maximizing the restricted log-likelihood using the *lme* function from "nlme" (version 3.1-150) R (version 4.0.3) package. Models were checked and validated by i) plotting the fitted vs. random residuals, ii) inspecting the distribution of the random residuals, and iii) inspecting the distribution of the random residuals across blistering status categories. Variance proportions explained by models was calculated using the "r.squaredGLMM" function from the MuMIn (version 1.43.17) R package. Microbial traits were included as responses variable, and subject to sqrt-root transformation when distributions were skewed.

Indicator species analysis was applied in indicspecies using the "r.g." function [7] and $10^5$ permutations. Four separate indicator analyses were conducted, including, i) the control group (n = 868) versus the patient group (n = 1,451) for all body site sampling locations, ii) the corresponding sampling sites in controls (n = 334) versus the combined patient perilesional (n = 415) and contralateral sites (n = 407) and , iii) the control group versus the patient group for all non-lesional sites (antecubital fossa = 363, upper back = 383, and forehead = 398), and iv) perilesional (n = 415) versus contralateral (n = 407) sampling sites only within the patient group. For each indicator analysis, ASVs were defined within each sample with a minimum threshold of 2%. The ASVs which met these criteria for all samples were then included in the analysis. Significant indicator ASVs were selected after correction of *p*-values for multiple testing using the Benjamini and Hochberg method [6].

To further evaluate indicator species results, we performed Random Forest classification and regression analyses using randomForest (version 4-6-14; [8]. Two sample sets, i) all controls (n = 868) and patient (n = 1,451) samples for all body sites, and ii) corresponding (n = 334) control- and patient perilesional (n = 415), and contralateral (n = 407) sites, were assessed using two distinct models: i) significant ASVs within each indicator species analysis, and ii) ASVs classified to the *Staphylococcus* genus whose abundance met the 2% minimum read threshold (as described above). Models were constructed with n = $10^5$ trees, while the "mtry" parameter was set for each model.

To disentangle the effects of disease, individual features, and sampling variation on the identified indicator ASVs and diversity measures, we constructed linear models that included the following explanatory variables: disease status (control or BP patient), body site, study center, sex, age, sequencing run, and DNA extraction round. Two respective models were constructed, including one for sites rarely affected by BP (antecubital fossa, upper back, and forehead) and one for perilesional, contralateral, and control corresponding sites. Response variables included significant indicator ASVs that met statistical significance in the indicator analyses. When necessary, square root transformations were applied on response variables to correct for a non-normal distribution. Models were checked and validated by examining the distribution of residuals, plotting the fitted and residual values, and examining the distribution of residuals for included explanatory variables. We reported the $R^2$ and the Beta coefficient value, which expresses the directionality of the effect (the degree of change in the response variable).

Moreover, we performed a non-parametric multivariable analysis of variance using distance matrices (PERMANOVA) using the “adonis” function with 1,000 permutations and a partial constrained principal coordinate analysis (PCoA) of beta diversity measures using the “capscale” function in vegan [9]. Disease status, body site, and study center were included as constraints, whereas sex, age, sequencing run, and extraction round were defined as conditions. The significance of models, axes, and terms were assessed using the “anova.cca” function with 1,000 permutations. These analyses were performed respectively for each of the two models described above.

Pairwise correlations among best indicator ASVs were evaluated using Spearman’s correlations, and *p*-values were corrected for multiple testing using Benjamini and Hochberg method [6].

### SUPPLEMENTARY MATERIAL RESULTS

#### Alpha diversity

We examined the influence of disease status, body site, sex, age, sequencing run, DNA extraction round, and study center on alpha diversity from patients and controls for sites rarely affected by BP (forehead, upper back, and antecubital fossa). We find that the body site and study center significantly correlate with Shannon diversity ($F_{39,1123} = 6.69$; $R^2_{adj} = 0.1603$, $p < 0.001$). Analysis of sum of squares reveals that body site explains a considerable fraction of variance (5.57%), whereas study center accounts for a comparably smaller fraction of the variance (2.5%). Interestingly, sex explains the highest percentage of variance (6.6%), while disease status is not significant. Similar to Shannon diversity, differences in Chao1 index for these same body sites significantly correlate with study center (4.9%), sequencing run (4.5%), and to a lesser extent, body site (0.5%) and sex (1.19%), while disease status is not significant ($F_{39,1123} = 4.15$; $R^2_{adj} = 0.096$; $p < 0.001$). Thus, overall, our statistical framework further emphasizes that the effects of disease on alpha diversity do not extend to non-lesional sites, and that rather the signal of skin biogeography predominates at these sites.

Next, to examine whether perilesional and contralateral sampled from the same individual (spatial correlation), as well as whether multiple samples taken from patients and controls are confounding, we fitted a linear mixed effects where blistering status (perilesional or contralateral, or control corresponding) was defined as the fixed effect and individual ID was defined as a random term. Blistering status significantly correlates with Shannon and Chao1 diversity in perilesional, contralateral, and corresponding sites (anova lme, blistering status $p < 0.0001$, variance explained $R^2= 8.61\%$; anova lme, blistering status $p= 0.0016$, variance explained $R^2= 1.69\%$, respectively.)

Additionally, we examined the potential confounding effect of spatial correlation in the sites rarely involved (i.e., forehead, antecubital fossa, and upper back) in BP. We find disease status does not correlate with Shannon and Chao1 diversity at forehead, antecubital fossa, and upper back (anova lme, disease status p=0.749, body site p <0.0001, $R^2$ = 5.33%; anova lme, disease status p=0.9336, body site p = 0.001, $R^2$= 0.61%, respectively.) In summary, we find similar effects of disease status and blistering status when accounting for spatial correlation.

**Beta diversity**

We examined the sites rarely affected by BP (forehead, upper back, and antecubital fossa) using PERMANOVA analysis applied to the Bray-Curtis index as measure of beta diversity, and find that disease status, body site, and study center influence bacterial community structure, in addition to sex, age, sequencing run, and extraction round (Supplementary Methods; adonis: disease status $R^2$ = 0.0109, body site $R^2$ = 0.044, study center $R^2$= 0.03, sex $R^2$ = 0.02, age $R^2$ = 0.001, sequencing run $R^2$ = 0.01, and extraction round $R^2$=0.01; $p < 0.05$; 1,000 permutations).

We analyzed beta diversity to examine potential differences in community structure for perilesional, contralateral, and control corresponding body sites. Associations not reported in main text include: disease status $R^2$=0.0212, blistering status $R^2$ = 0.0029, study center $R^2$ = 0.0341, sex $R^2$ = 0.013, age $R^2$ = 0.001, sequencing run $R^2$ = 0.02, and extraction round $R^2$ = 0.016 (Supplementary Method, adonis: $p < 0.05$; 1,000 permutations). We find significant associations with disease (i.e., control corresponding versus patient) and blistering status (i.e., control corresponding versus perilesional and contralateral sites), which explain 2.12% and 0.29% of the variance, respectively (adonis: disease status $R^2$=0.02; blistering status $R^2$ = 0.003; $p < 0.001$). In this model, **s**tudy center explains 3.41% of the variance (adonis: $R^2$ = 0.03; $p < 0.001$), suggesting environmental or geographical effects.

Next, to further examine the relationship between beta diversity and the BPDAI severity score, and to account for spatial correlation, we calculated a Bray-Curtis index using mean ASV values from perilesional and contralateral sites separately (n = 210). Congruent with the PERMANOVA results presented in main text, we find BPDAI and sampling center are significantly associated with beta diversity, suggesting perhaps that only the most severely affected patients are seen at university study centers. Results reported in tables below.

**Table 1.** Bray-Curtis index results using mean ASV values from patient perilesional sites

| Perilesional sites | $R^2$ | p value |
|---|---|---|
| **Sex** | 0.01773 | 0.002997 |
| **Age** | 0.00457 | 0.392607 |

| | | |
|---|---|---|
| **BPDAI (disease severity score)** | 0.01336 | 0.001998 |
| **Sampling Center** | 0.08163 | 0.000999 |
| **Run** | 0.04676 | 0.401598 |
| **Extraction round** | 0.04567 | 0.134865 |

**Table 2.** Bray-Curtis index results using mean ASV values from patient contralateral sites

| Contralateral sites | $R^2$ | p value |
|---|---|---|
| **Sex** | 0.01980 | 0.000999 |
| **Age** | 0.00532 | 0.192807 |
| **BPDAI (disease severity score)** | 0.01726 | 0.000999 |
| **Sampling Center** | 0.07761 | 0.000999 |
| **Run** | 0.04999 | 0.106893 |
| **Extraction round** | 0.04348 | 207792 |

**Correction of spatial correlation for the best ASV indicators**

When accounting for spatial correlation (by including "individual" as a random term), we find similar results in terms of significance and proportions of variance explained, except for ASV1_*C. acnes,* which is no longer significantly affected by blistering status (perilesional, contralateral, or corresponding).

**Table 3.** ANOVA LME results for best ASV indicators after accounting for spatial correlation

| ASV | Model | p value | Variance explained $R^2$ |
|---|---|---|---|
| **ASV_1** ***C. acnes*** | ANOVA LME | 0.103 | 0.69% |
| **ASV_2** ***S. aureus*** | ANOVA LME | < 0.0001 | 12.42% |
| **ASV_4** ***C. tuberculostearicum*** | ANOVA LME | < 0.0001 | 3.73% |
| **ASV_5** ***S. hominis*** | ANOVA LME | 4e-04 | 1.5% |

### REFERECNCES

[7] De Caceres M, Legendre P. Associations between species and groups of sites: indices and statistical inference. Ecology 2009.

[8] Liaw A, Wiener M. Classification and Regression by randomForest. Newsl R Project 2002;2:5.

[9] Oksanen J, Blanchet FG, Friendly M, Kindt R, Legendre P, McGlinn D, et al. Community Ecology Package. 2005.

### SUPPLEMENTARY FIGURES

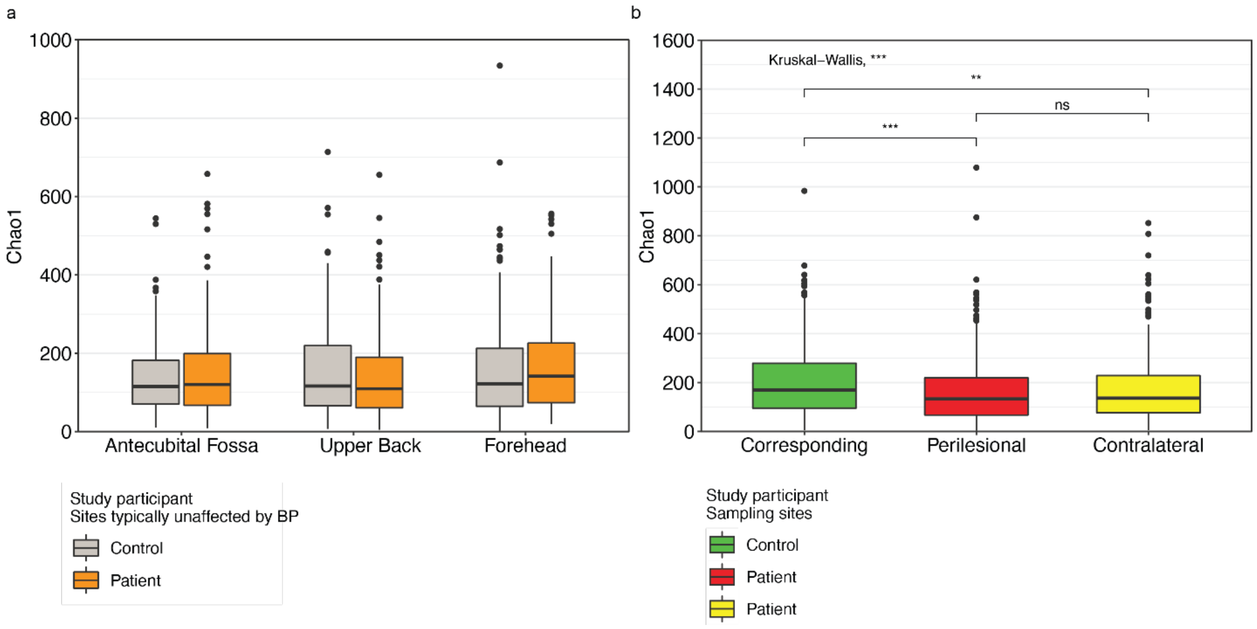

**Supplement Figure S1**. **S1a**. Chao-1 index for sites rarely affected by BP (forehead, upper back, and antecubital fossa) between patients and controls. **S1b.** Chao-1 index for the perilesional and contralateral sites of the patient group and corresponding site in controls. For box plots: Boxes represent interquartile range between first and third quartiles; horizontal line defines the median. Whiskers represent lowest and highest values. Kruskal-Wallis test applied to analyze site variation. If an overall significant difference was observed, a pairwise Wilcox test was performed; p-values adjusted using the Benjamini-Hochberg method. Significance represented by: * ≤ 0.05; ** ≤ 0.01; *** ≤ 0.001; **** ≤ 0.0001. Supplementary Table S6 reports summary statistics.

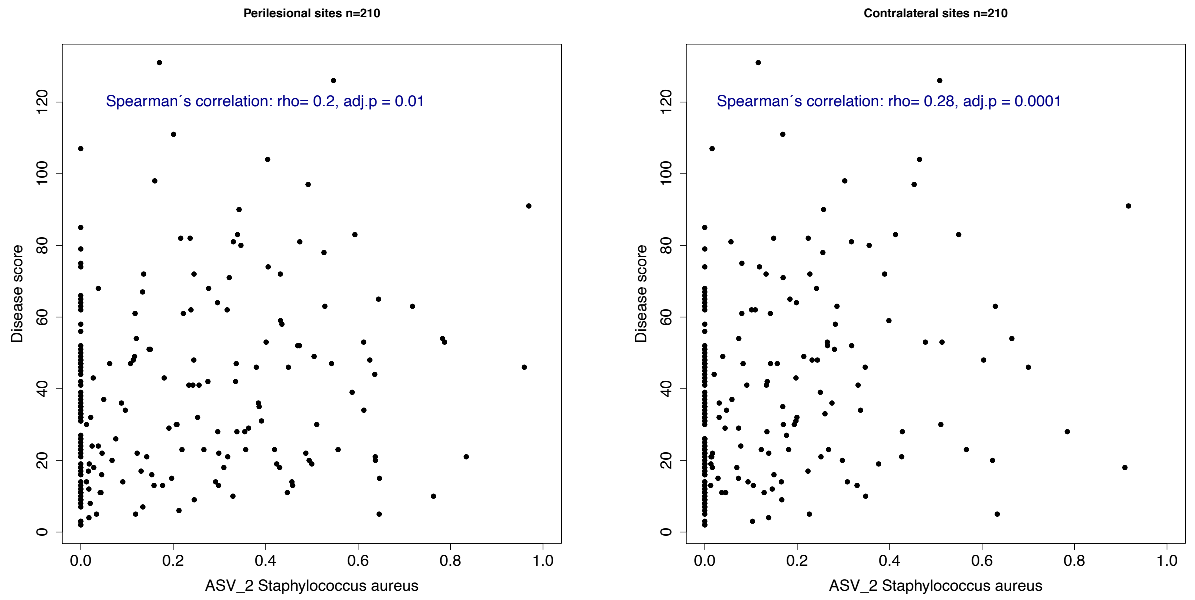

**Supplement Figure S2.** *S. aureus* correlation with BPDAI (only activity accounted).

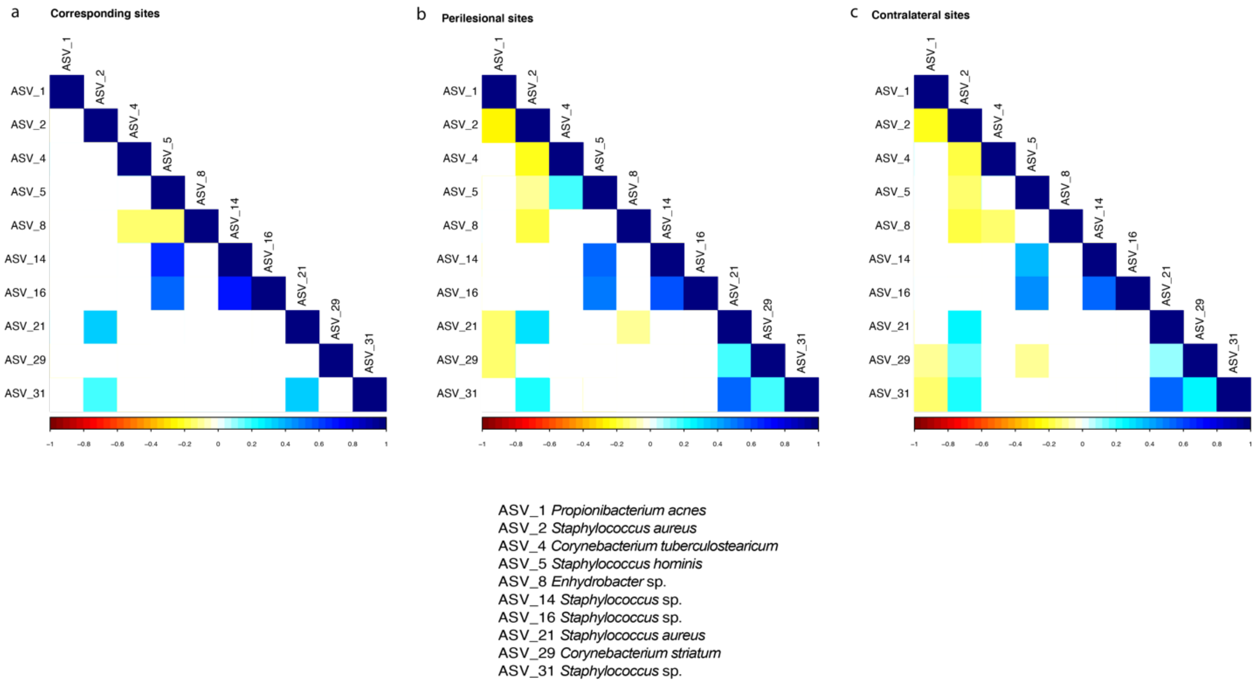

**Supplementary Figure S3**. Spearman's correlation matrix between **S3a**. Indicator ASVs and control corresponding sampling sites, **S3b.** Indicator ASVs and patient perilesional sampling sites, and **S3c.** Indicator ASVs patient contralateral sampling sites. Blank box indicates $p > 0.05$ after correction for multiple testing.

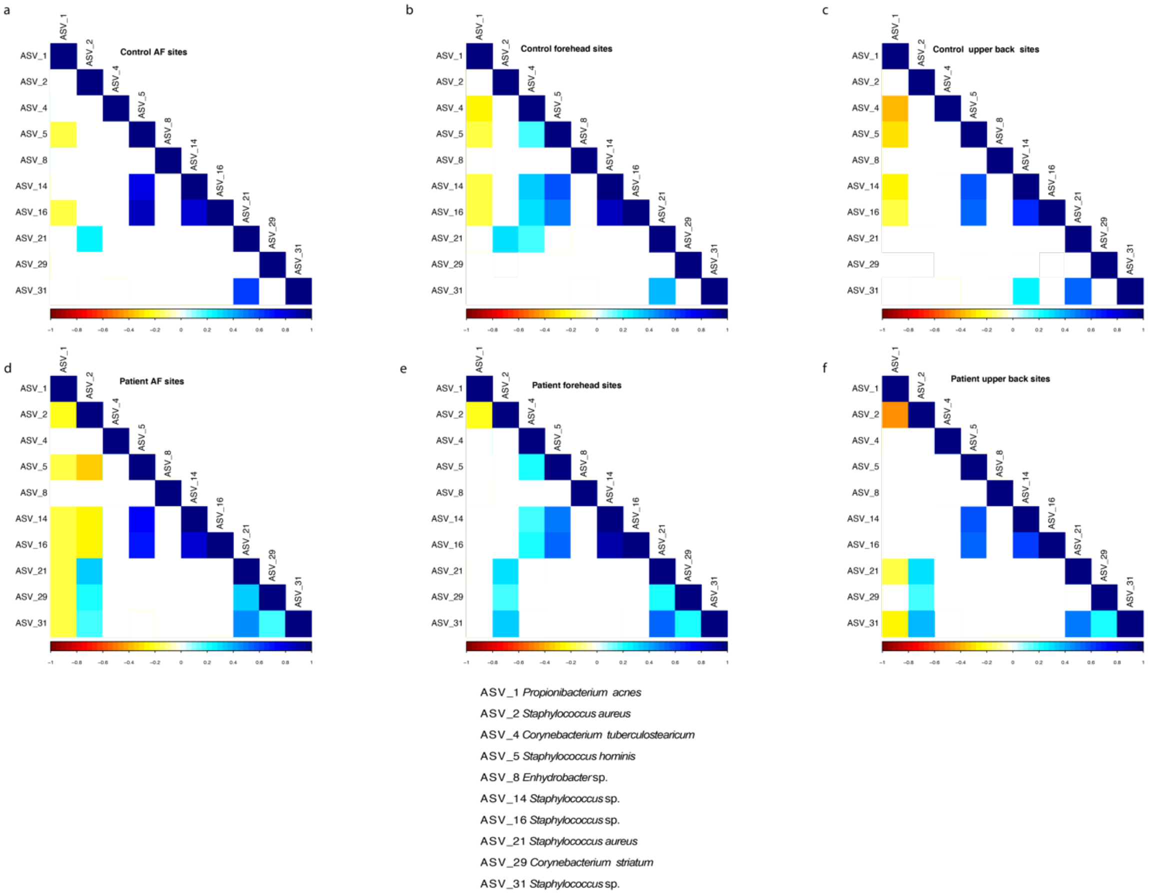

**Supplementary Figure S4.** Spearman's correlation matrix between, **S4a.** Indicator ASVs and antecubital fossa (AF) sites from matched controls, **S4b.** Indicator ASVs and forehead sites from matched controls, **S4c.** Indicator ASVs and upper back sites from matched controls, **S4d**. Indicator ASVs and antecubital fossa (AF) sites from patients with BP, **S4e.** Indicator ASVs and forehead sites from patients with BP, **S4f.** Indicator ASVs and upper back sites from patients with BP.
Blank box indicates $p > 0.05$ after correction for multiple testing.

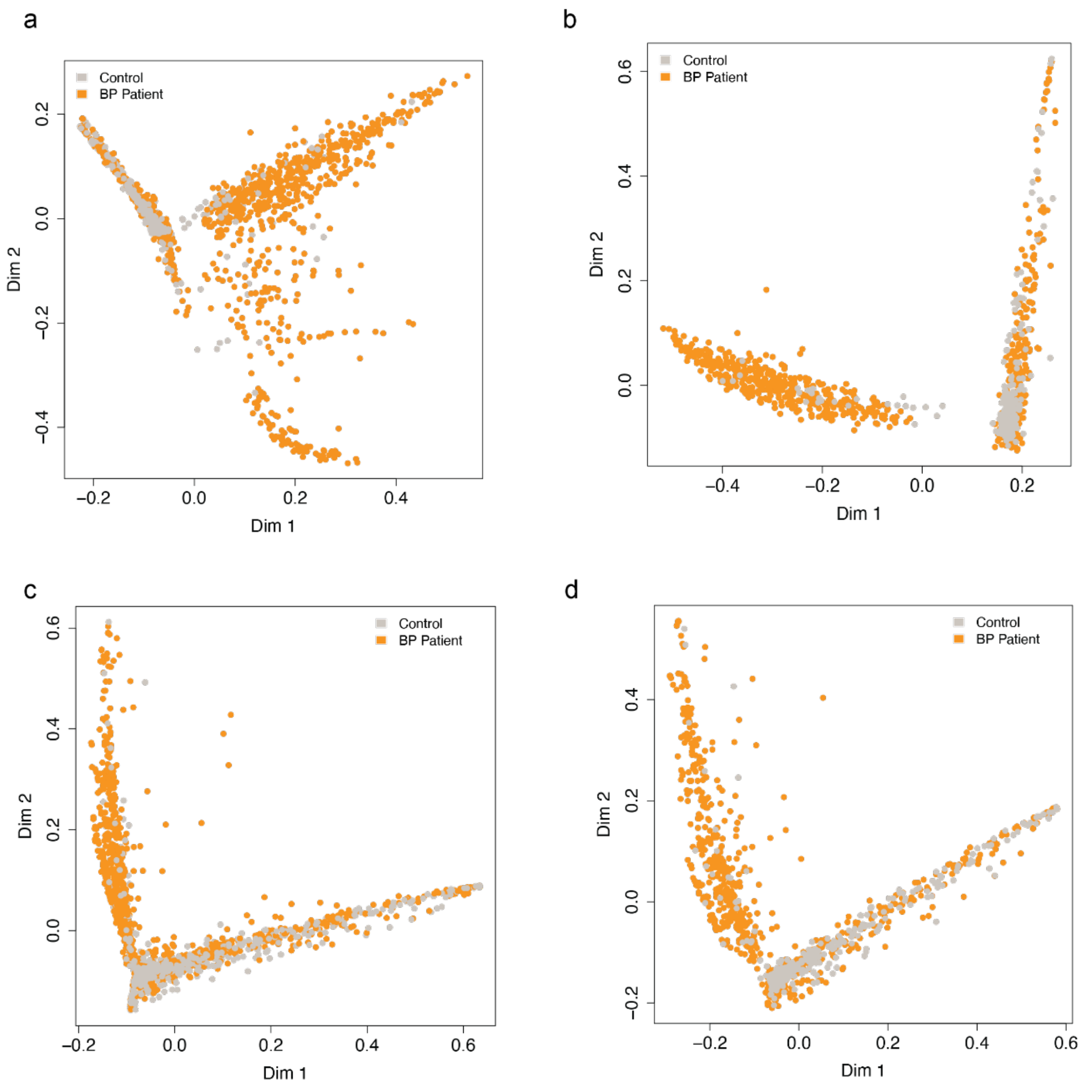

**Supplementary Figure S5.** Multidimensional plot of the proximity matrix by randomForest analysis for the four tested models (Methods). **S5a.** Controls versus patients with BP for all identified indicators (n = 370) for all sampling sites (mtry = 15); (849/868 controls, and 1443/1451 patients with BP, mean classification accuracy 99%) **S5b.** Controls versus patients with BP for all identified indicators (n =3 70) for only control corresponding and patient perilesional and contralateral sampling sites (mtry = 18); (324/334 controls, and 822/822 patients with BP, mean classification accuracy 99.15%). **S5c.** controls versus patients with BP for all sampling sites, but limited indicators to *Staphylococcus* ASVs with an abundance >2% within each sample; (mtry = 52, 790/868 controls, and 1,446/1,451 patients with BP, mean classification accuracy 96.4%). **S5d.** controls versus patients with BP for control corresponding, patient perilesional, and patient contralateral sampling sites and limited to *Staphylococcus* ASV indicators with an abundance >2% within each sample (mtry = 62, controls 294/334, and 819/822 patients with BP, 96.2%).
